# Evaluation of a phenotypic, point-of-care solution for the detection and quantitative antibiotic susceptibility testing for lower Urinary Tract Infection

**DOI:** 10.1101/2021.12.17.21267412

**Authors:** R. Turner, R. Kirkby, E. Meader, J. Wain

## Abstract

**Background:** Urinary tract infections (UTIs) are one of the most common bacterial infections seen in primary care. The current standard for diagnosis is microbiological culture and antibiotic sensitivity testing of a mid-stream urine sample; however, this technique is costly, labour intensive and typically takes 2-3 days to yield a result.

**Study design and Objective:** This is a nonexperimental cross-sectional study. The aim of this study was to evaluate the efficacy of *U-treat*, a bioluminescent approach for rapid detection of bacteriuria and *quantitative* determination of the antimicrobial susceptibility profiles of uropathogens in clinical urine specimens - in under an hour.

**Method:** The evaluation was carried out in two UK-based Medical Centres using urine samples from patients presenting with symptoms of a UTI (n=249). The *U-treat* technology is a two test, two reagent process. Test 1 detects the presence of a bacterial UTI > 10^4^ bacteria/mL (5-10 minutes). Test 2 produces *quantitative* antibiotic susceptibility (<50 minutes). Only urine samples testing positive for bacteria in Test 1 underwent Test 2 (n=82). *U-treat* results were compared retrospectively against reference laboratory culture and sensitivity findings. The influence of the technology on patient treatment outcomes was also analysed.

**Results:** Relative to reference laboratory analysis, Test 1 showed a sensitivity of 97.1% and specificity of 92.0%. (PPV: 89.3%; NPV: 97.8%). Test 2 produced an overall sensitivity (measurement of true susceptibility) of 94.1% (Predictive value: 96%) and an overall specificity (measurement of true resistance) of 90.5% (Predictive value 86.4%). Analysis of treatment data demonstrated that had the physicians had access to *U-treat* results at the point of care, the percentage of patients treated successfully would have risen from 68.3% to 92.7%.

**Conclusion:** *U-treat* represents the first technology, world-wide, capable of providing UTI treatment data to physicians at the point of care, in less than 60 minutes.

## Introduction

Approximately 150 million urinary tract infections (UTIs) occur annually worldwide^1^. UTIs are one of the most common clinical bacterial infections in women, accounting for nearly 25% of all infections. Around 50– 60% of women will develop UTIs in their lifetimes and one in three will have at least one UTI necessitating antibiotic treatment by age 24^2^. Suspected UTIs account for up to 3% of all GP visits. In England alone, this adds up to around million consultations, and costs the NHS more than £316 million in GP time^3^. The Medical Technology Group has found that in 2012/14 the NHS spent £434 million on treating 184,000 emergency admissions caused by a urinary tract infection. This is an average per patient cost of £2,361. The cost of UTIs globally exceeds $6billion in direct health-care expenditures^4^.

More than 95% of lower UTI or cystitis is associated with a single pathogen. The predominant causal organism is *Escherichia coli* 5. General features of cystitis include dysuria, urgency and a sensation of incomplete bladder emptying, lower abdominal pain and haematuria. The current UK guidance for GP management of a suspected UTI case varies depending on patient age, symptoms and the results of dipstick analysis. However, the gold standard for confirmatory diagnosis remains culture and antibiotic sensitivity analysis of a midstream, urine specimen referred to an external clinical laboratory. GPs can find it challenging to diagnose UTIs; the 2-3 day delay in receiving external culture and sensitivity results combined with patient pressure and the risk of complication, frequently leads to unwarranted precautionary antibiotic prescriptions^6^.

UTIs account for about 15% of antibiotics prescribed in primary-care and up to 60% of patients treated with antibiotics do not have a microbiologically proven UTI^7,8^. This inappropriate use of antibiotics is an important driving factor of antimicrobial resistance (AMR) development. More than 700,000 people die of drug-resistant infections every year, and this figure is expected to reach 10 million by 2050 (United Nations meeting on AMR, 2016). Increasing AMR complicates UTI treatment by increasing patient morbidity, costs of reassessment and re-treatment and use of broader spectrum antibiotics. A 2016 survey evaluating the point of care test (POCT) needs of UK GPs found the number one POCT which clinicians felt would help them in their diagnostic decision-making was that for UTIs^9^.

In this study we evaluate, using a nonexperimental cross-sectional study design, the efficacy of *U-treat*, a novel phenotypic point-of-care technology which utilises the principles of ATP-bioluminescence to detect a bacterial UTI and determine quantitative antibiotic susceptibility, within in an hour. We hypothesise that such a tool would provide practitioners with real time specific guidance on the need for antibiotic treatment and would directly help the implementation of the NICE guidelines on antibiotic stewardship.

## Materials and Methods

This evaluation was carried out at two Medical Centres by staff with no knowledge of the clinical information nor the reference method results. Human urine samples were collected from mid-stream patient samples presenting with symptoms typical of UTI using aseptic vacuum technique, and aliquoted prior to being forwarded to Norfolk & Norwich University Hospital (NNUH) Microbiology for automated microscopy and, where appropriate, urine culture and sensitivity analysis. Patient and Public Involvement were not incorporated in this study.

An application was granted for Ethical Approval of a Health-Related Research Project by the University of East Anglia Faculty of Medical and Health Sciences Research Ethics Committee. Study design and applications were carried out according to an agreed Service Evaluation protocol. Patient confidentiality was maintained at all times using an anonymised alpha-numeric coding system of patient urine samples, processed after routine standard microbiology had been completed. Results of Test 1 and Test 2 were compared retrospectively with NNUH Microbiology records on-line by authorised Medical Centre personnel. Only the NHS staff member had access to NHS patient data. In addition to comparison of Test 1 and Test 2 with reference laboratory analysis, treatment data were collected on an ongoing basis from retrospective results comparison with Practice staff (Fig.1).

**Figure 1:**
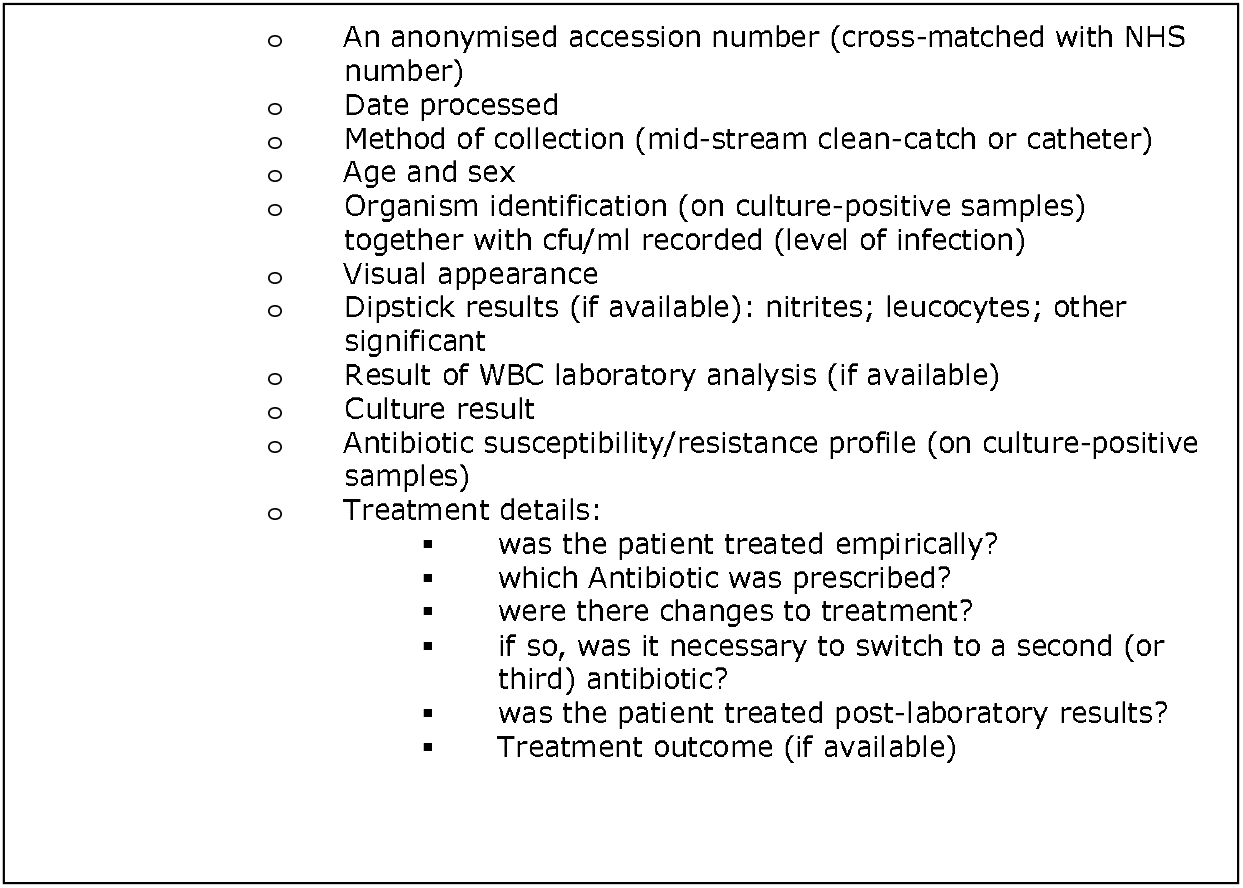
Data capture.

The technology utilises a two-reagent, two-test system. Reagent 1 contains a mix of detergents, nutrient broth and buffered enzymes designed to permeabilise non-bacterial cells within the target sample, releasing host cell ATP. This host cell ATP, together with free ATP, is dephosphorylated, making it unavailable for the stage two reaction. Following incubation at 37°C, a second reagent contained in a delivery device is ejected into the reagent 1 mix. Reagent 2 contains a mix of buffered luciferase/luciferin and detergent, designed to permeabilise bacterial cell membranes; the bacterial ATP released is captured by the luciferase/luciferin mix, producing a light signal output directly proportional to the level of bacteria present^10,11^. Relative Light Units (RLU’s) are captured on a bespoke luminometer.

Test 1 detects bacteriuria. The signal output cut-off for Test 1 was calculated at 5000RLUs, from values determined on culture-negative and culture-positive urine samples. Test 2 quantitatively measures the responses of uropathogens to antibiotics at the end of a 30-minute incubation period; the signal outputs directly compares the effect of each antibiotic on the bacterial population with a control tube that does not contain an antibiotic. Antibiotics were formulated for use in the *U-Treat* assay to reflect breakpoint concentrations [EUCAST: Clinical breakpoints and dosing of antibiotics]. The resistance/susceptibility cut-off was based on a calculated 25% kill rate, from values determined on culture-positive samples. Positive and Negative Controls were included to ensure all equipment and reagents performed as required. The luminometer software provides visual data in the form of a bar chart, that allowed the operator to easily determine the most effective antibiotic to treat the detected bacterial infection (Fig. 2).

**Figure 2:**
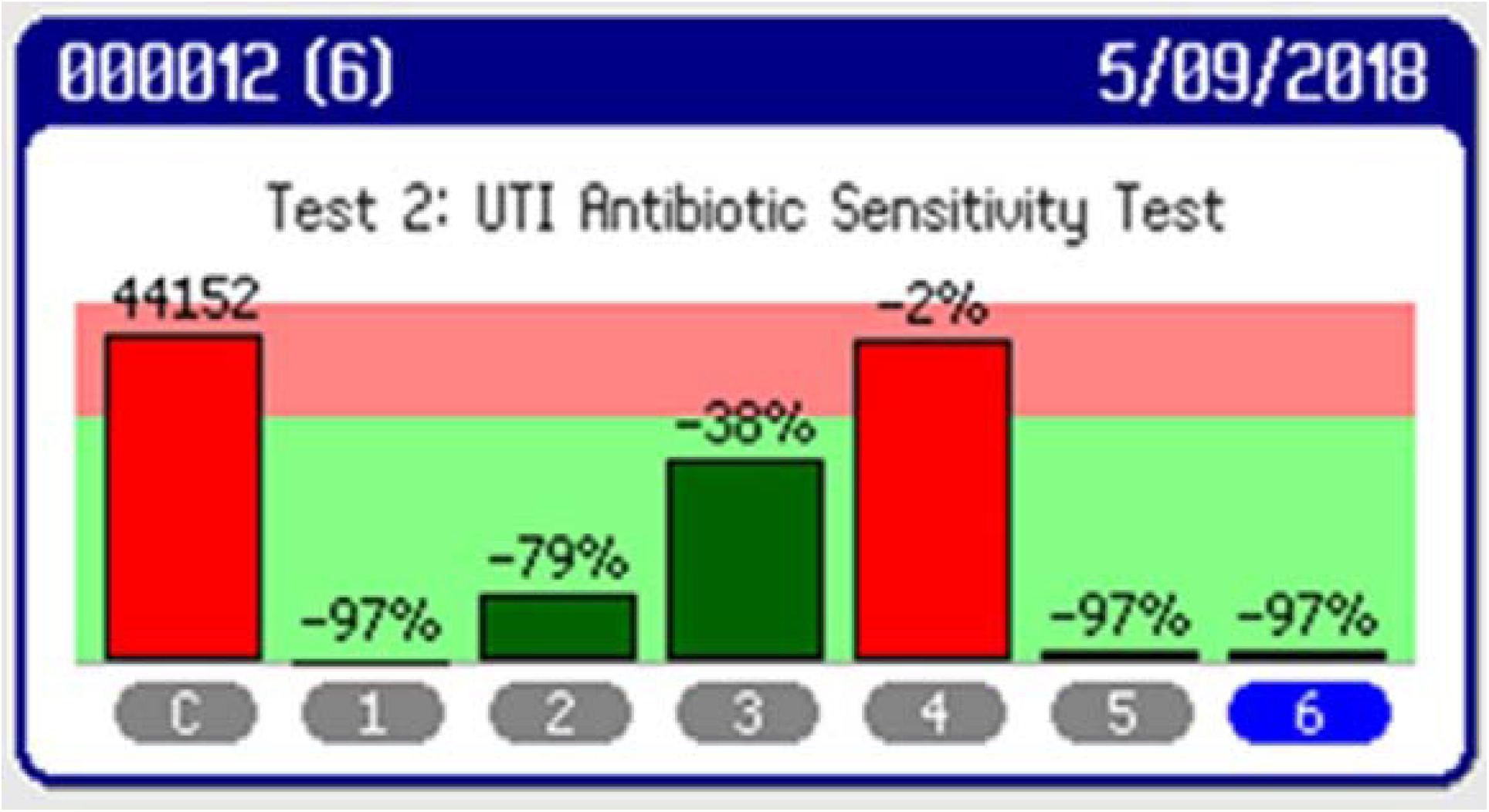
Lumini screen shot – Test 2 AS profile.

### Data analysis – comparison to reference method

Sensitivity and specificity were calculated by comparison with reported results from the Norfolk and Norwich University Hospital (NNUH) Microbiology Laboratory; the laboratory provides an acreditated service to clinicians. Urine samples submitted from local Medical Centres for UTI, are processed using an algorithm that utilises a fully automated flow cytometer (Sysmex UF500) together with traditional culture on chromogenic media for detection of UTI. Samples determined to be positive for UTI are then processed utilising mass spectroscopy for microbial identification and antibiotic susceptibility systems (Vitek MS). The NNUH Microbiology staff had access to routine clinical information but were unaware that *U-Treat* was also being used to analyse these specimens.

### Treatment outcomes

Treatment outcomes were categorised as: Successful when:

1. a patient determined as not suffering with a UTI, based on symptoms, was not prescribed with an antibiotic
2. a patient determined as likely suffering with a UTI, based on symptoms and history, was prescribed an effective antibiotic at first presentation

Unsuccessful when:

1. a culture-negative patient was inappropriately prescribed with an antibiotic
2. a culture-positive patient was prescribed an antibiotic that the UTI did not respond to, resulting in a switch to a second antibiotic, based on reference laboratory results

## Results

### Test 1 – Bacterial Detection

A total of 270 urines from patients presenting to their respective GP with symptoms of a cystitis: dysuria, urgency and a sensation of incomplete bladder emptying and lower abdominal pain, were used in this study. A total of 249 results were compared with gold standard reference laboratory findings. The NNUH Microbiology laboratory reported 146 as being culture - negative for UTI and 103 as being culture-positive for UTI; results are presented in Table 1. For the remaining 21 samples the reference laboratory returned no result because the samples contained contaminants. When tested using U-Treat Test 1, one was negative and twenty were positive. For Test 2, these twenty positive samples gave no susceptibility result and so the patients would have been asked to provide a new specimen. As no test Test 2 result (antimicrobial susceptibility) was obtained for these samples they were removed from the statistical analysis.

**Table 1:** Summary of Test 1 results (UTI detection)

| Test 1 | True Positive | False Negative | True Negative | False Positive |
| --- | --- | --- | --- | --- |
| n= 249 |  |  |  |  |
| Med Centre 1 | 67 | 1 | 96 | 8 |
| Med Centre 2 | 33 | 2 | 38 | 4 |
| Combined sites | 100 | 3 | 134 | 12 |

**False positives and false negatives = non-agreement with laboratory findings**

Sensitivity was calculated to be 97.1% and Specificity 91.8% (PPV: 89.3%; NPV: 97.8%)

When broken down by Health centre data indicates that the test was performed with equal consistency (Table 2).

**Table 2:** Statistical summary of Test 1 results (UTI detection)

| Test 1 | GP Med | GP Med | Combined |
| --- | --- | --- | --- |
| n=249 | Centre 1 | Centre 2 |  |
| % Sensitivity (95% CL) | 98.5 (92.1 to 99.7) | 94.3 (81.4 to 98.4) | 97.1 (91.8 to 99.0) |
| % Specificity (95% CL) | 92.3 (85.6 to 96.1) | 90.5 (77.9 to 96.2) | 91.8 (86.2 to 95.2) |
| POSITIVE likelihood ratio | 12.8 | 9.9 | 11.8 |
| NEGATIVE likelihood ratio | 0.02 | 0.06 | 0.03 |
| Disease prevalence | 39.5 | 45.5 | 41.4 |
| POSITIVE predictive value | 89.3 | 89.2 | 89.3 |
| NEGATIVE predictive value | 99 | 95 | 97.8 |

### Test 2 - Antibiotic Susceptibility

Results on samples that were considered as “contaminants” by the reference clinical laboratory were not included in the comparative analysis of Test 2 results. A total of 82 Test 2 antibiotic profiles were compared with reference laboratory results reported to each Medical Centre.

Overall Sensitivity (measurement of true susceptibility): 94.1% (90.0 to 96.6, 95% confidence limits). Predictive value (all antibiotics) calculated as 96%.

Overall Specificity (measurement of true resistance): 90.5% (82.3 – 95.1, 95% confidence limits). Predictive value (all antibiotics) calculated as 86.4%.

### Treatment outcomes

Statistical percentages obtained for each parameter were similar for both Medical Centres. These included: the percentage of patients treated empirically; the percentage of treatments that were changed after empirical treatment; the percentage of UTI-negative patients (confirmed by laboratory culture) that were treated unnecessarily with an antibiotic and the percentage treated following receipt of laboratory results (Table 4).

**Table 3:** Summary of Test 2 results (Antibiotic Susceptibility)

| Combined Medical Centres | True Susceptible | False Resistant | False Susceptible | True Resistant |
| --- | --- | --- | --- | --- |
| NIT | 58 | 3 | 1 | 3 |
| AMX | 23 | 3 | 2 | 35 |
| AMC | 17 | 1 | 0 | 16 |
| CLX | 57 | 1 | 3 | 0 |
| TMP | 37 | 4 | 2 | 22 |
| ALL antibiotics | 192 | 12 | 8 | 76 |

**Table 4:** Summary of treatment outcomes - combined Medical Centres 1&2

|  | Med Centre | Med Centre 2 | Combined |
| --- | --- | --- | --- |
| Parameter | 1 | 2 | 1&2 |
| n (Patients treated) | 52 | 30 | 82 |
| No. treated empirically | 30 | 17 | 47 |
| % treated empirically | 57.7% | 56.7% | 57.3% |
| No. empirical correctly treated | 12 | 9 | 21 |
| % empirical correctly treated | 40% | 52.9% | 44.7% |
| Treatment change post empirical treatment | 9 | 4 | 13 |
| % changed post empirical treatment | 30% | 23.5% | 27.7% |
| No. UTI Negs treated empirically | 9 | 4 | 13 |
| % UTI-Negs treated empirically | 30% | 23.5% | 27.7% |
| No. inappropriate treatment | 18 | 8 | 26 |
| No. treated post-lab results | 22 | 13 | 35 |
| % treated post-lab results | 42.3% | 43.3% | 42.7% |
| Total correct treatment | 34 | 22 | 56 |
| % correct treatment | 65.4 | 73.3 | 68.3 |

**Table 5:** Summary of treatment outcome if U-Treat technology had been adopted

|  | Med Centre | Med Centre 2 | Combined |
| --- | --- | --- | --- |
| Parameter | 1 | 2 | 1&2 |
| n (Patients treated) | 52 | 30 | 82 |
| No. treated empirically | 30 | 17 | 47 |
| T&T would have correctly treated: | 27 | 17 | 44 |
| % T&T would have correctly treated | 90% | 100% | 93.60% |
| No. inappropriate treatment (T&T) | 3 | 0 | 3 |
| % inappropriate empirical treatment | 10% | 0% | 6.4% |
| % correct empirical treatment | 90% | 100% | 93.6% |
| No. UTI Negs treated empirically | 9 | 4 | 13 |
| T&T outcome would have been: | 0 | 0 | 0 |
| No. treated post-lab results | 22 | 13 | 35 |
| T&T outcome /agreement with clinical lab | 20 | 12 | 33 |
| Total correct treatment (T&T) | 47 | 29 | 76 |
| % correct treatment (T&T) | 90.4% | 96.7% | 92.7% |

Treatment outcomes utilising the current algorithm (Public Health England, UTI Treatment Guidelines, UK Government documents for Health Professionals 2018) resulted in less than half (44.7%) of empirically treated patients being correctly treated (Table 4). For thirteen UTI-negative cases (27.7%) where antibiotics were prescribed, *U-treat* Test 1 produced a negative signal output for all thirteen and would have prevented the use of antibiotics in these patients.

## Discussion

The *U-treat* technology could improve antibiotic stewardship and patient outcomes for the treatment of UTI in general practice. The technology involves measurement of a light signal output, produced when bacterial adenosine triphosphate (ATP) binds to luciferin-luciferase^12^. ATP bioluminescence has been applied in the food industry and in hygiene monitoring to detect the presence of bacteria for over 30 years^10,13,14^. However, utilising ATP bioluminescence to detect and manage antibiotic susceptibility at the point of care provides a novel tool that reflects the dynamics of bacterial metabolic activity; either indicating presence of bacteria or the level of reactivity to the presence of antibiotics that disrupt bacterial metabolic activity. The application of this technology provides a biochemical solution to a microbiological problem – the method reliably detects infections and produces data on which to base antibiotic treatment decisions, in a timeframe suitable to the point-of-care setting.

Uropathogens adhere, colonise and adapt to the nutritionally limited bladder environment. They evade immune surveillance and persist and can disseminate the urinary tract^15^.The presence of leucocytes, immune-response-associated cells and cast material are common in the urine of UTI patients^16^. These non-bacterial or host cells contain ATP. Urine also contains free ATP from the breakdown of host cells. Host cell and free ATP form a background of “noise”. Reagent 1 eliminates somatic cell-derived ATP and free ATP, without affecting bacterial cell membranes. Thereby, the final reaction selectively detects bacterial ATP and the associated light signal output is directly proportional to the density of bacteria present^10,11^.

Results obtained utilising a POCT technology should have an acceptably high sensitivity and specificity according to WHO ASSURED guidelines. In this study, results from combined sites indicate an acceptable Test 1 performance, producing a sensitivity of 97.1% and specificity of 92%. Data generated comparing *U-treat* Test 2 with reported clinical laboratory antibiotic susceptibility results, indicate clinically acceptable performance. Test 2 produced an overall sensitivity (measurement of true susceptibility) of 94% and an overall Specificity (measurement of true resistance) of 91%^17,18,19^.

A 2020 retrospective study exploring the diagnostic work-up and treatment of UTIs in an out-of-hours clinic found 74% of all patients visiting the clinic with UTI symptoms received antibiotics^6^. In this study, 57% of patients going to their GP for UTI symptoms were treated empirically with antibiotics, of which 27.7% were culture-negative and 27.7% received an inappropriate antibiotic. A major advantage of *U-Treat*, as illustrated by this evaluation, is the generation of rapid, *quantitative* susceptibility data. Had GPs had access to the results of the *U-treat* assay at point-of-care, over 90% of those treated empirically would have received the correct treatment; none of the culture-negative patients would have been prescribed antibiotic treatment and, of the cases where treatment was switched to a second antibiotic post laboratory results, *U-Treat* would have identified the correct antibiotic at the time of presentation in all of the patients. By reducing unnecessary antibiotic prescriptions and ensuring appropriate treatment at the time of presentation, *U-treat* has the potential to improve antibiotic stewardship as well as improve patient outcomes.

Of the 43% of patients who awaited laboratory culture and sensitivity results prior to treatment, *U-treat* test results correlated with laboratory results in 94% of cases. Using the current PHE algorithm, 31.7% of patients in this study received inappropriate treatment. Had both Medical Centres had access to the U-Treat technology 92.7% of the patients would have received appropriate treatment, reducing inappropriate therapy to <10%.

Potential limitations of this study include the removal of ‘contaminant samples’ and sample size. Suspected contaminant sample results (a total of 21 reported) were removed by NHS personnel, as the external laboratory was unable to definitively say the sample was a ‘true negative’. Contaminant samples could potentially interfere with *U-treat* outcomes. This evaluation included urine from a total of 249 patients presenting to their respective GP with symptoms of a cystitis - dysuria, urgency and a sensation of incomplete bladder emptying and lower abdominal pain. The results were compared with gold standard reference laboratory findings and consisted 103 culture-positive and 146 culture-negative samples. Resource constraints would not allow for an extended evaluation beyond this number of subjects but we believe that these preliminary data are encouraging.

Point-of-care testing is defined as diagnostic testing that is performed at or near to the site of the patient with the result leading to a potential change in the care of that patient. POCTs provide advantages over existing laboratory-based tests, due to their rapidity, allowing cost-effective and decentralised diagnosis of a wide range of infectious diseases and public health related threats^20^. Several point of care tests for UTI detection have been developed and commercialised, including culture-based and enzymatic assays, however all have their limitations^7,21,22^. According to EUCAST and CLSI guidelines, reliable antibiotic susceptibility testing may benefit more from phenotypic testing, i.e., a dynamic test assessing whether the microorganism grows in the presence of the antibiotic. Phenotypic methods work regardless of the resistance mechanism and give answers to the practical question of whether an antibiotic is likely to be effective at the dose used in the therapy^23^. *U-treat* is a phenotypic technology that, in addition, provides *quantitative* susceptibility data to allow a physician to make a prescription choice.

Given that 47% of GPs within the UK have expressed a need for a more accurate testing to aid the diagnosis of a UTI a PoC UT test would met a perceived clinical need. The data generated in this trial indicates the *U-Treat* technology would be of benefit in detection and treatment of UTIs at the point of care and represents the first technology, world-wide, able to do this in less than one hour. *U-treat* could, prospectively, obviate the need for empirical prescribing, decrease the external laboratory load of urine specimens and their associated costs, reduce the number of GP visits and hospital admissions secondary to mismanaged UTIs and curb the development of antimicrobial resistance ^17^. More research is required to look at the health economics and clinical utility of introducing such technology within the UK and globally.

## Data Availability

All data collected for this study are available via the University of East Anglia Library. PhD thesis Ronald Turner.

## Patents

Rapid Determination of Bacterial Susceptibility to an Antibiotic at The Point Of Care - filed in USA (granted August 2018), Europe (granted June 2019) and Canada (in process).

## Acknowledgments

We thank the following for support during the project.

Dr James Fisher & Colleagues, Acle Medical Centre, Norwich

Dr Nick Morton, Bowthorpe & Trinity Medical Centre, Norwich

Dr George Savva, UEA and Quadram Institute, Norwich

Dr Heather Felgate, UEA and Quadram Institute, Norwich

and Integrated Technologies Ltd, Kent

## Author’s contributions

RT Was involved with all aspects of the project and wrote the first draft of the manuscript. RK Checked validity of the results, helped with the original draft and edited the manuscript. EM Helped design the study, supervised the clinical work and edited the final manuscript. JW helped with the original draft of the manuscript, prepared the final draft for submission and supervised the non-clinical aspects of the work.

## Patient and Public Involvement statement

This is a laboratory based study and although informal enquiries were made to understand the process of GP referral from the side of the resource constraints meant it was not possible to formally involve patients in the design, or conduct, or reporting, or dissemination plans of our research.

## Data sharing

All data collected for this study are available via the University of East Anglia Library. PhD thesis Ronald Turner. We will disseminate data through sharing with the UK’s Longitude Prize Committee and there was no sponsor for this study.

